# Data Heterogeneity in Federated Learning with Electronic Health Records: Case Studies of Risk Prediction for Acute Kidney Injury and Sepsis Diseases in Critical Care

**DOI:** 10.1101/2022.08.30.22279382

**Authors:** Suraj Rajendran, Zhenxing Xu, Weishen Pan, Arnab Ghosh, Fei Wang

## Abstract

With the wider availability of healthcare data such as Electronic Health Records (EHR), more and more data-driven based approaches have been proposed to improve the quality of care delivery. Predictive modeling, which aims at building computational models for predicting clinical risk, is a popular research topic in healthcare analytics. However, concerns about privacy of healthcare data may hinder the development of effective predictive models that are generalizable because this often requires rich diverse data from multiple clinical institutions. Recently, federated learning (FL) has demonstrated promise in addressing this concern. However, data heterogeneity from different local participating sites may affect prediction performance. Exploring such heterogeneity of data sources would aid in building accurate risk prediction models in FL. Due to acute kidney injury (AKI) and sepsis’ high prevalence among patients admitted to intensive care units (ICU), the early prediction of these conditions based on AI is an important topic in critical care medicine. In this study, we take AKI and sepsis onset risk prediction in ICU as two examples to explore the impact of data heterogeneity in the FL framework for risk prediction using EHR data across multiple hospitals. In particular, we built predictive models based on local, pooled, and FL frameworks. The local framework only used data from each site itself. The pooled framework combined data from all sites. In the FL framework, each local site did not have access to other sites’ data. A model was trained locally and its parameters were shared to a central aggregator, which was used to update the federated model’s weights and then subsequently, shared with each site. We found models built within a FL framework outperformed local counterparts. Then, we analyzed variable importance discrepancies across sites and frameworks. Finally, we explored potential sources of the heterogeneity within the EHR data. The different distributions of demographic profiles, medication use, and site information contributed to data heterogeneity.

**Author Summary:** The availability of a large amount of healthcare data such as Electronic Health Records (EHR) and advances of artificial intelligence (AI) techniques provides opportunities to build predictive models for disease risk prediction. Due to the sensitive nature of healthcare data, it is challenging to collect the data together from different hospitals and train a unified model on the combined data. Recent federated learning (FL) demonstrates promise in addressing the fragmented healthcare data sources with privacy-preservation. However, data heterogeneity in the FL framework may influence prediction performance. Exploring the heterogeneity of data sources would contribute to building accurate disease risk prediction models in FL. In this study, we take acute kidney injury (AKI) and sepsis prediction in intensive care units (ICU) as two examples to explore the effects of data heterogeneity in the FL framework for disease risk prediction using EHR data across multiple hospital sites. In particular, multiple predictive models were built based on local, pooled, and FL frameworks. The local framework only used data from each site itself. The pooled framework combined data from all sites. In the FL framework, each local site did not have access to other sites’ data. We found models built within a FL framework outperformed local counterparts. Then, we analyzed variable importance discrepancies across sites and frameworks. Finally, we explored potential sources of the heterogeneity within EHR data. The different distributions of demographic profiles, medication use, site information such as the type of ICU at admission contributed to data heterogeneity.

## Introduction

Acute kidney injury (AKI) and sepsis are two types of potentially life-threatening clinical conditions X, that complicate treatment, clinical trajectories, and potentially worsen outcomes of a significant number of hospitalized or intensive care unit (ICU)-patients [1-2]. For patients with AKI or sepsis, morbidity and mortality are usually higher than patients without AKI or sepsis, with as much as a sevenfold increased mortality risk, regardless of type of ICU (for example, medical, surgical, or cardiac) [3-4]. Moreover, healthcare utilization within the ICU is often higher for patients with these conditions. For example, patients with AKI and sepsis often require hemodialysis, inotropic support, or mechanical ventilation [5]. Therefore, early prediction of AKI or sepsis risk in critical care settings can facilitate early interventions that are likely to provide benefit, including aggressive treatment with fluid resuscitation and antimicrobials that may improve patient outcomes [6].

Recently, due to wider availability of electronic health record (EHR) data and advances in artificial intelligence (AI), machine learning (ML) based disease risk prediction has attracted more attention in the ICU setting [7]. Building an accurate and generalizable disease risk prediction model requires a large amount of data from a diverse patient population. Collecting the data together from different hospitals and constructing a unified risk prediction model on the combined data can lead to better prediction performance X. Moreover, using multiple hospitals or sites data over single institution data can add to the generalizability of ML models [8]. A recent study has shown that creating more generalizable models can increase algorithmic fairness, yet many published models lack this generalizability across geographic locations and demographics [9]. However, due to the highly sensitive nature of EHR in terms of protected health information (PHI) of patients, aggregating multiple institutions’ data all together is challenging.

More recently, federated learning (FL) has emerged as a promising strategy on building ML models with fragmented sensitive data [10]. FL is one mechanism of training ML models across multiple decentralized sites holding local data samples without exchanging them [11]. It builds a central aggregator to obtain global ML model’s parameters by iteratively exchanging model parameters from local ML models. However, data heterogeneity in the FL framework may affect prediction performance [12]. For example, different hospitals have different populations, which may have a high degree of variability in the patient treatment, such as different medications they administer and different procedures they conduct. Investigating the heterogeneity of data sources will aid in building an accurate disease risk prediction model in FL.

Although FL methods have been proposed, to our knowledge there have been no published studies that systematically investigate the data heterogeneity in FL for AKI and sepsis risk prediction. The aim of this study is to investigate the effects of data heterogeneity in the FL framework for disease risk prediction using EHR data across multiple hospital sites. In particular, we built predictive models based on local, pooled, and FL frameworks, and analyzed variable importance discrepancies across sites and its ramifications. The local framework only used data from each site itself. The pooled framework combined data from all sites. In the federated model framework, each local site does not have access to other sites’ data. They trained a model locally and shared parameters to a central aggregator, which was used to update federated model parameters and shared with each site. Finally, we explore potential sources of the heterogeneity within EHR data. The overall workflow of our study is shown in Fig 1.

**Fig 1.**
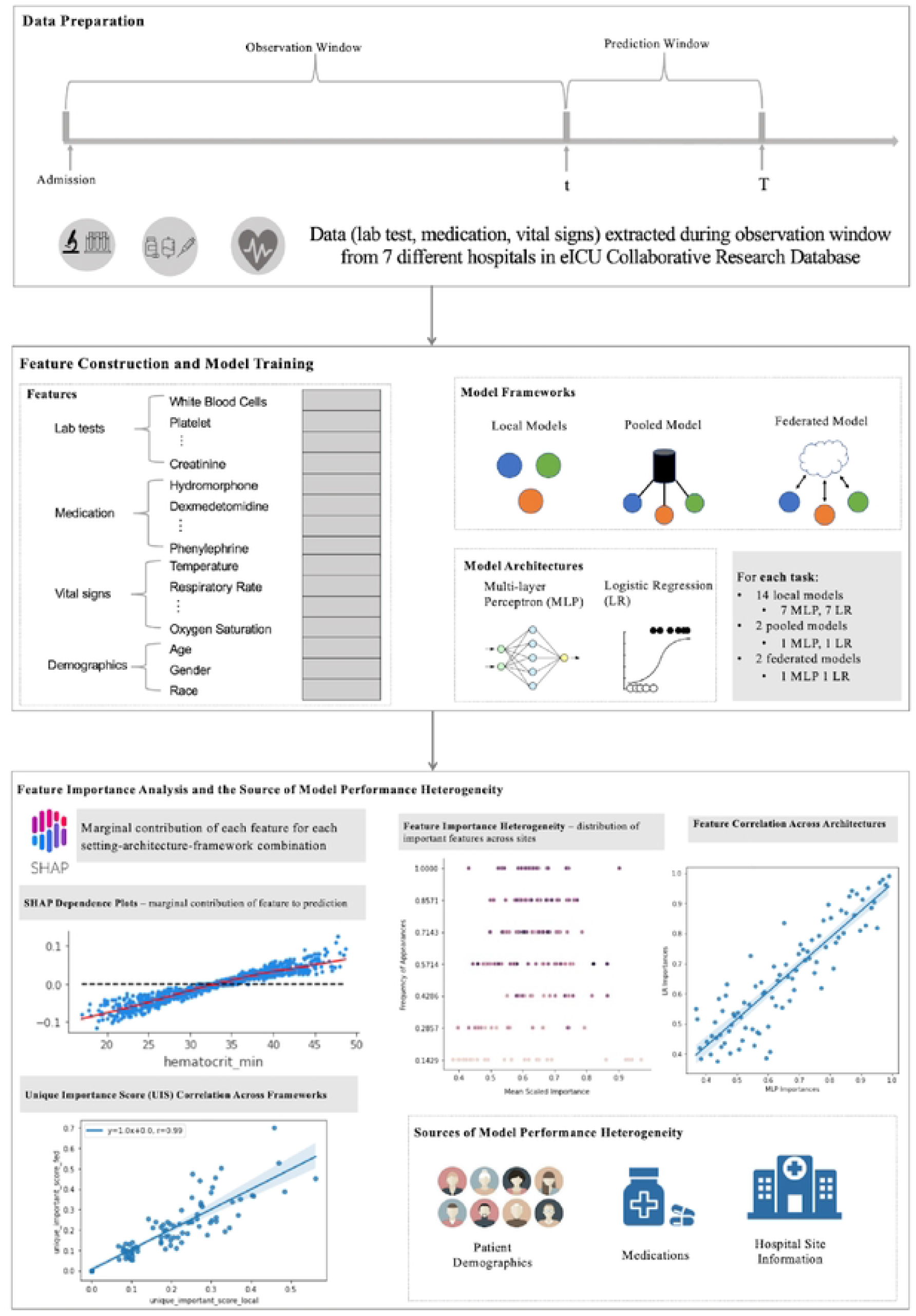
The framework of the study. In Data Preparation, different types of data including lab test, medication, vital signs, and demographic arc extracted during the observation window, which are used to build patients’ profiles to predict whether or not they would suffer from acute kidney injury or sepsis in the prediction window. In Feature Construction and Model Training, individual features from lab test, medication, vital signs, and demographic were obtained to build a predictive model based on three frameworks including local, pooled, and federated frameworks. In each framework, two common model architectures including logistic regression and multi-layer perceptron were used. In Feature Importance Analysis and Source of Model Performance Heterogeneity, feature importance heterogeneity, feature correlation across mode architectures and frameworks, and sources of model performance heterogeneity were explored.

## Results

### Development of AKI and sepsis prediction models in local sites

Data for 21,796 AKI patients and 22,0082 sepsis patients at 7 hospitals were extracted from the eICU collaborative research database, following inclusion and exclusion criteria denoted in the “Methods” section. All patients shared 354 unique variables which included lab tests, vital signs, demographics, and medications. AKI patients were labeled both within a 24h and 48h observation window, leading to two settings for AKI prediction. For sepsis, we labeled patient data in accordance with Sepsis-3 clinical criteria. We predicted whether patients would suffer from sepsis 6 hours prior to onset, onset point included. Within the observation window, lab tests and vital sign information were aggregated through several statistics (minimum, maximum, first, and last values) into several new features. Three model frameworks were designed including local, pooled, and federated model architectures. The details of each model architecture were described in the “Methods” section. For each framework, two model architectures including the multilayer perceptron (MLP) and logistic regression (LR) were explored. Processed data and scripts used for analyses are also available at https://github.com/surajraj99/Data-Heterogeneity-in-Federated-Learning.

Fig 2 illustrates the LR and MLP performance, measured by area-under-receiver-operator-curve (AUC), on both AKI 24h and 48h settings. Sepsis prediction setting results can be found in Supplemental Information and Supplemental Fig 6. We observed:

- When using local model framework: AKI 24h LR models performed within the range of 0.680 - 0.809, whereas MLP models performed within a range of 0.677 - 0.821. Similarly, AKI 48h LR models performed within the range of 0.680 - 0.809, whereas MLP models performed within a range of 0.673 - 0.800. Sepsis LR models’ performances ranged between 0.771 - 0.834 across sites, whereas MLP models’ performances ranged between 0.772 - 0.829. The LR and MLP models performed similarly across all prediction tasks.
- When using pooled model framework: AKI 24h LR models performed within the range of 0.672 - 0.742, whereas MLP models performed within a range of 0.78 - 0.827. Similarly, pooled AKI 48h LR models performed within the range of 0.683 - 0.744, whereas MLP models performed within a range of 0.686 - 0.755. Pooled sepsis LR models’ performances ranged between 0.731 - 0.800 across sites, whereas MLP models’ performances ranged between 0.732 - 0.793. LR pooled models showed more consistent performances to local model counterparts in comparison to MLP pooled models.
- When using federated model framework: AKI 24h LR models performed within the range of 0.742 - 0.834, whereas MLP models performed within a range of 0.732 - 0.839. Similarly, AKI 48h LR models performed within the range of 0.722 - 0.835, whereas MLP models performed within a range of 0.72 - 0.833. Federated sepsis LR models’ performances ranged between 0.833 - 0.862 across sites, whereas MLP models’ performances ranged between 0.823 - 0.861.
- Generally, the federated model outperformed the local model and pooled model. The pooled models underperformed the local model.

**Fig 2.**
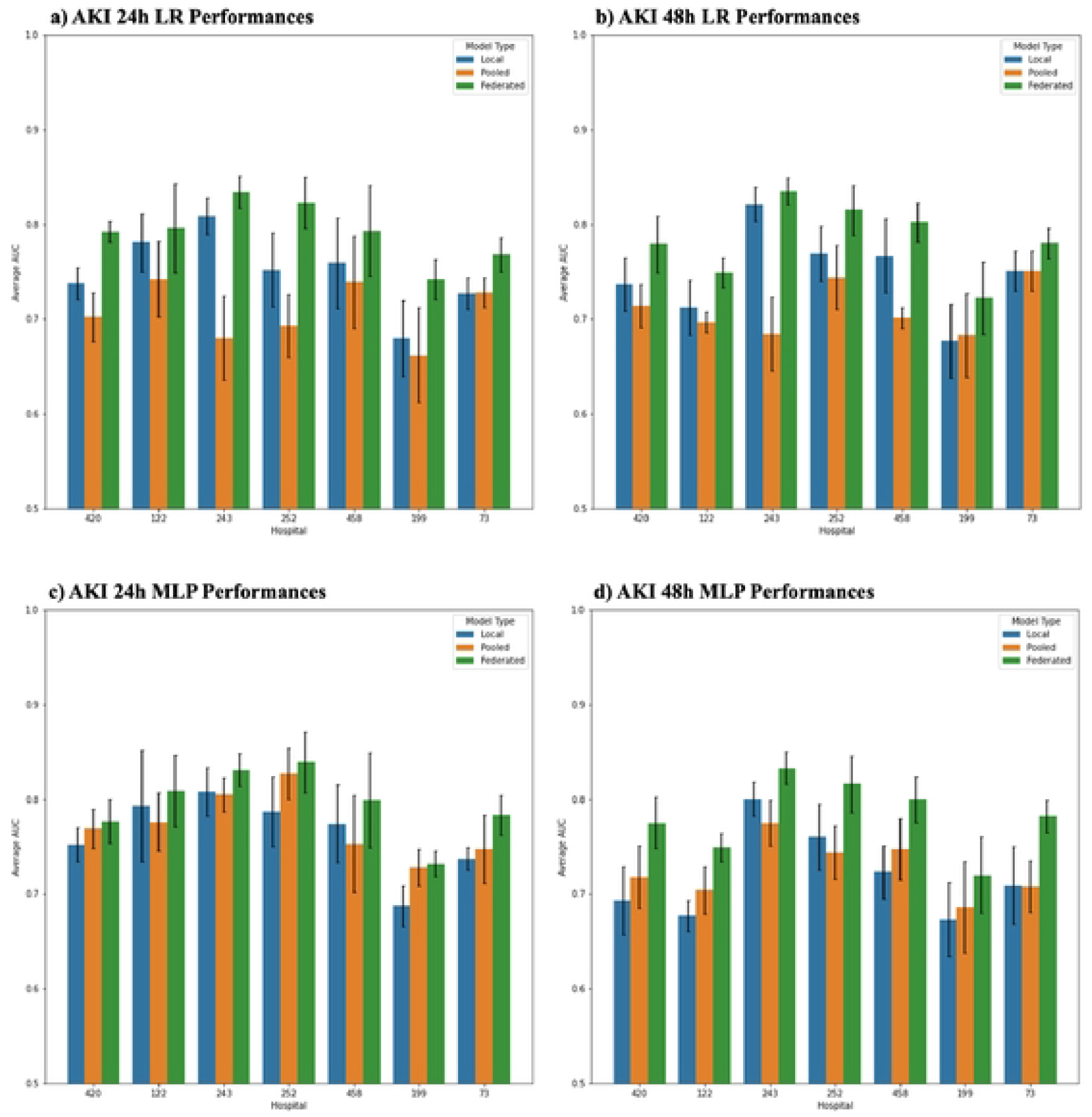
Area Under Receiver Operating Curve (AUROC) for AKI Setting. Each plot shows performances for AKI 24h and 48h prediction settings. Blue bars depict each local site’s model performance on their respective site lest data. Orange bars depict pooled model performance on each local site’s test data. Green bars depict federated model performance on each local site’s test data.

### Clinical interpretation of sepsis and AKI prediction models

Using Shapley Additive exPlanations (SHAP) values, we investigated the marginal effects of the features identified as predictive by each model. Fig 3 illustrates the marginal plots (SHAP dependence plots) for the top 10 most important features for each pooled model on the AKI prediction settings. Fig 4 shows the SHAP dependence plots for AKI setting federated models. Dependence plots for all local models are available in S4 Fig. Sepsis prediction results are available in the Supplemental Information and S7 Fig.

**Fig 3.**
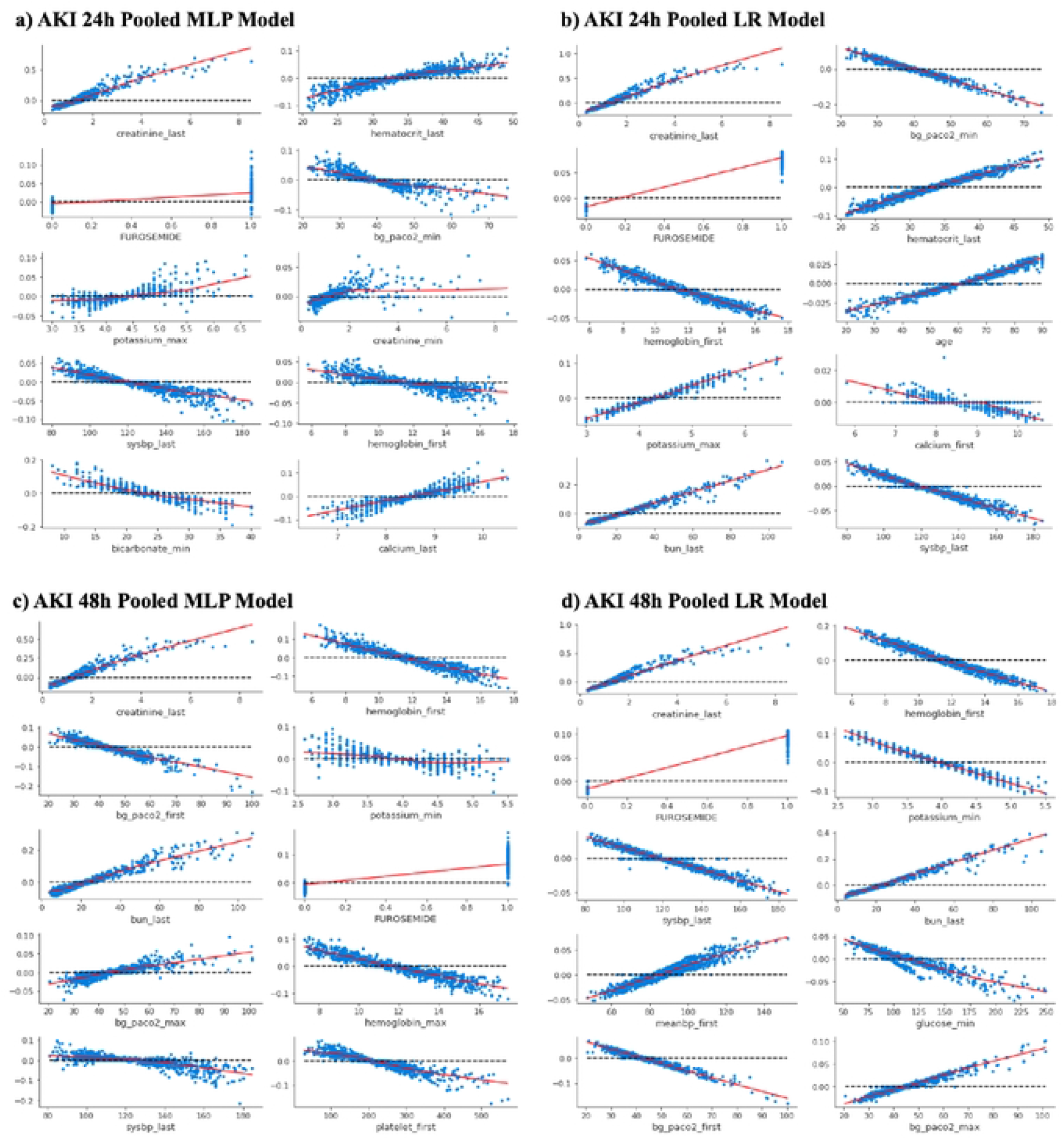
Shapley dependence plots for top 10 features for pooled models. Each panel shows the marginal effects of each of the most impactful features ranked among the top 10 for predicting AKI 24h or 48h using pooled models. The x-axis gives the raw values of each feature and the y-axis gives the logarithmic of estimated odds ratio (i.e.. the SHAP value) for sepsis. AKI 24h or AKI 48h. when a feature takes a certain value. Each dot represents the SHAP value of a sample. The LOWESS curve, used for smoother extrapolating across all the dots, is plotted in red for all panels, (a. c) show Shapley dependence plots for pooled MLP models and (b. d) show Shapley dependence plots for pooled LR models, (a. b) show plots for AKI 24h. and (c. d) show plots for AKI 48h.

**Fig 4.**
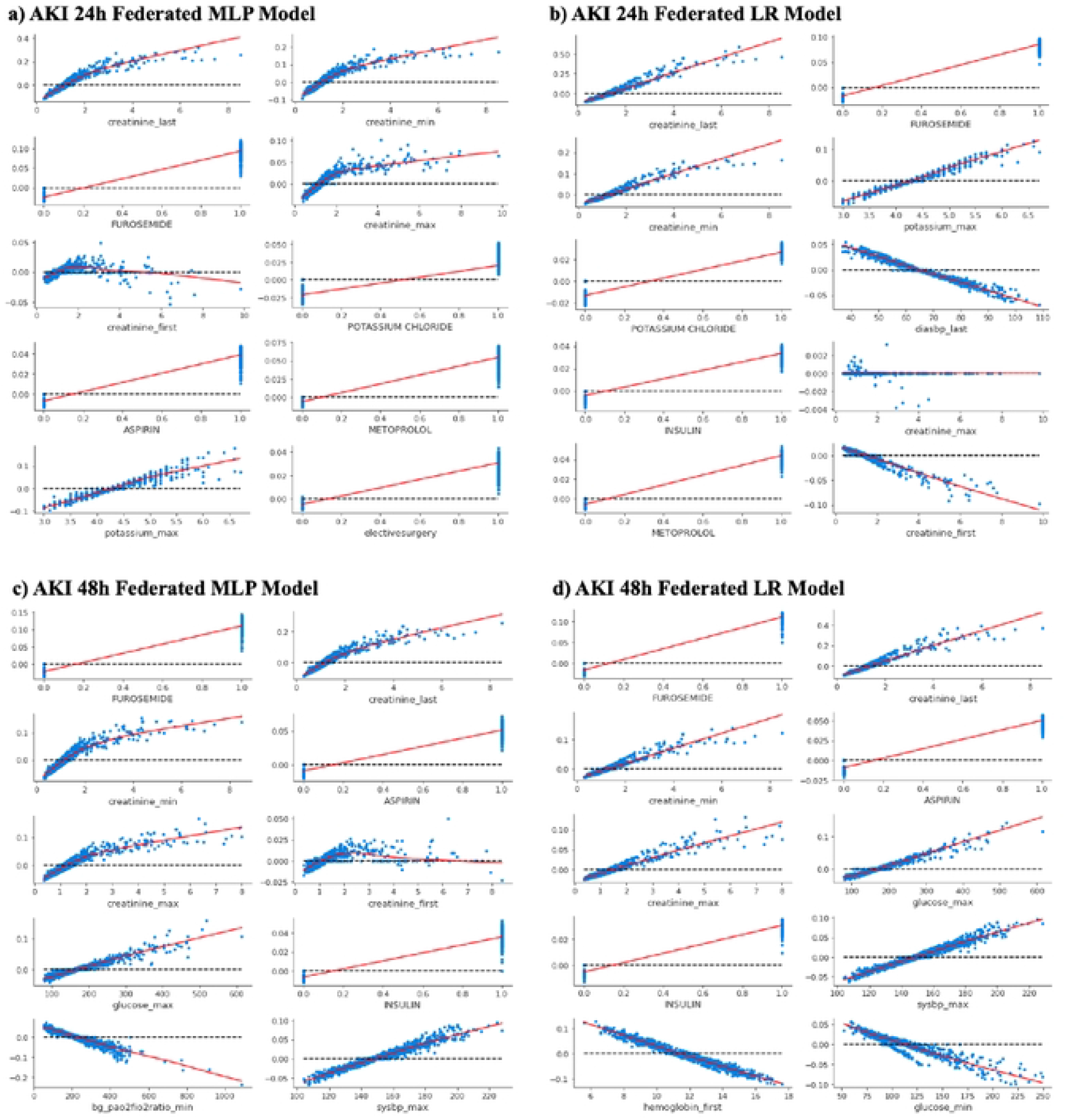
Shapley dependence plots for top 10 features for federated models. Each panel shows the marginal effects of each of the most impactful features ranked among the top 10 for predicting AKI 24h or 48h using federated models. The x-axis gives the raw values of each feature and the y-axis gives the logarithmic of estimated odds ratio (i.e., the SHAP value) for sepsis. AKI 24h or AKI 48h. when a feature takes a certain value. Each dot represents the SHAP value of a sample. The LOWESS curve. used for smoother extrapolating across all the dots, is plotted in red for all panels, (a. c) show Shipley dependence plots for federated MLP models and (b. d) show Shipley dependence plots for federated LR models, (a. b) show plots for AKI 24h. and (c. d) show plots for AKI 48h,

In the AKI 24h setting, the pooled MLP model identified last measured level of creatinine *(creatinine_last)*, last measured hematocrit level *(hematocrit_last)*, Furosemide, *bg_paco2_min*, maximum potassium level *(potassium_max)*, minimum creatinine levels *(creatinine_min)*, last measured systolic blood pressure *(sysbp_last), hemoglobin_first*, minimum bicarbonate level *(bicarbonate_min)*, and last measured calcium level *(calcium_last)* as the top 10 most important variables. All factors except Furosemide were lab tests and vital signs. The pooled LR model shared several important factors with the pooled MLP model, with the addition of age, first measured calcium level *(calcium_first)*, and last measured blood urea nitrogen level *(bun_last)*. Of particular note, in the pooled MLP model, *creatinine_last* of ~4 mg/dL is associated with an exp(0.4) = 1.5 fold increase in risk of AKI 24h. In the pooled LR model, *creatinine_last* shows a similarly strong relationship as the pooled MLP to AKI 24h risk. A *bun_last* measurement of ~60 mg/dL is associated with a exp(0.2) = 1.2 fold increase in risk of AKI 24h. In the pooled LR model, the risk of AKI 24h given administration of furosemide, is greater than AKI risk in the MLP model, with an odds ratio of exp(0.1) = 1.1.

In the AKI 48h setting, the pooled MLP model identified *creatinine_last, hemoglobin_first, bg_paco2_first, potassium_min, bun_last*, Furosemide, maximum partial pressure of carbon dioxide (*bg_paco2_max), hemoglobin_max, sysbp_last*, and first measured platelet count *(platlet_first)* as the top 10 most important variables. All factors except Furosemide were lab tests and vital signs. The pooled LR model shared several important factors with the pooled model, with the addition of the mean systolic and diastolic blood pressure *(meanbp_first)* and minimum glucose level *(glucose_min)*. Similar to the 24h setting, in the 48h pooled MLP model, *creatinine_last* of ~4 mg/dL is associated with an exp(0.4) = 1.5 fold increase in risk of AKI. A *bun_last* measurement of greater than ~25 mg/dL is associated with an increased risk of AKI 48h. In the pooled LR model, *creatinine_last* and *bun_last* show similarly strong relationships as the pooled MLP model. Furosemide is considered an important medication across all AKI settings and model architectures.

For the AKI 24h setting, the federated MLP and LR model consider more medications important than their respective pooled counterparts. Medications considered important by the federated MLP model include Furosemide, Potassium Chloride, Aspirin, and Metoprolol, whereas the federated LR model considered Insulin important as well. Interestingly, the federated MLP model considered the patient’s choice of elective surgery *(electivesurgery)* as an important feature, albeit a relatively small increase (exp(0.02) = 1.02 fold) in risk of AKI 24h. Like the 24h setting, the federated MLP and LR models of the AKI 48h setting considered more medications important than their respective pooled counterparts. Both the MLP and LR model consider administration of Aspirin and Insulin as important factors. The federated MLP for the 48h setting uniquely finds the minimum ratio of “partial pressure of oxygen” to “fractional inspired oxygen” *(bg_pao2fio2ratio_min)* and maximum level of glucose *(glucose_max)* as important factors. Local models shared numerous important factors with pooled and federated models, depicting similar relationships between feature value and risk of sepsis/AKI (S4 Fig).

### Source of prediction performance heterogeneity across model architectures, frameworks, and sites

To better understand differences in feature importances across hospital sites and model frameworks, we performed a qualitative analysis which looked at the most important variables selected by models and their prevalence across sites. Fig 5, 6, and 7 show features in relation to their importance rankings in AKI prediction models, where the y-axis is the proportion of sites that consider the feature as a top 100 feature (for the specific model architecture). For example, a feature that has a y-value of 1.0 is deemed important at all sites, whereas if a feature has a y-value of 0.1429 (1/7), it is only considered important at one site. The x-axis shows the importance ranking of the feature, averaged across the sites it is considered important (i.e. top 100) in (i.e., a feature is more important if it is closer to 1). Results for the sepsis prediction setting are available in the Supplemental Information and S8 Fig.

**Fig 5.**
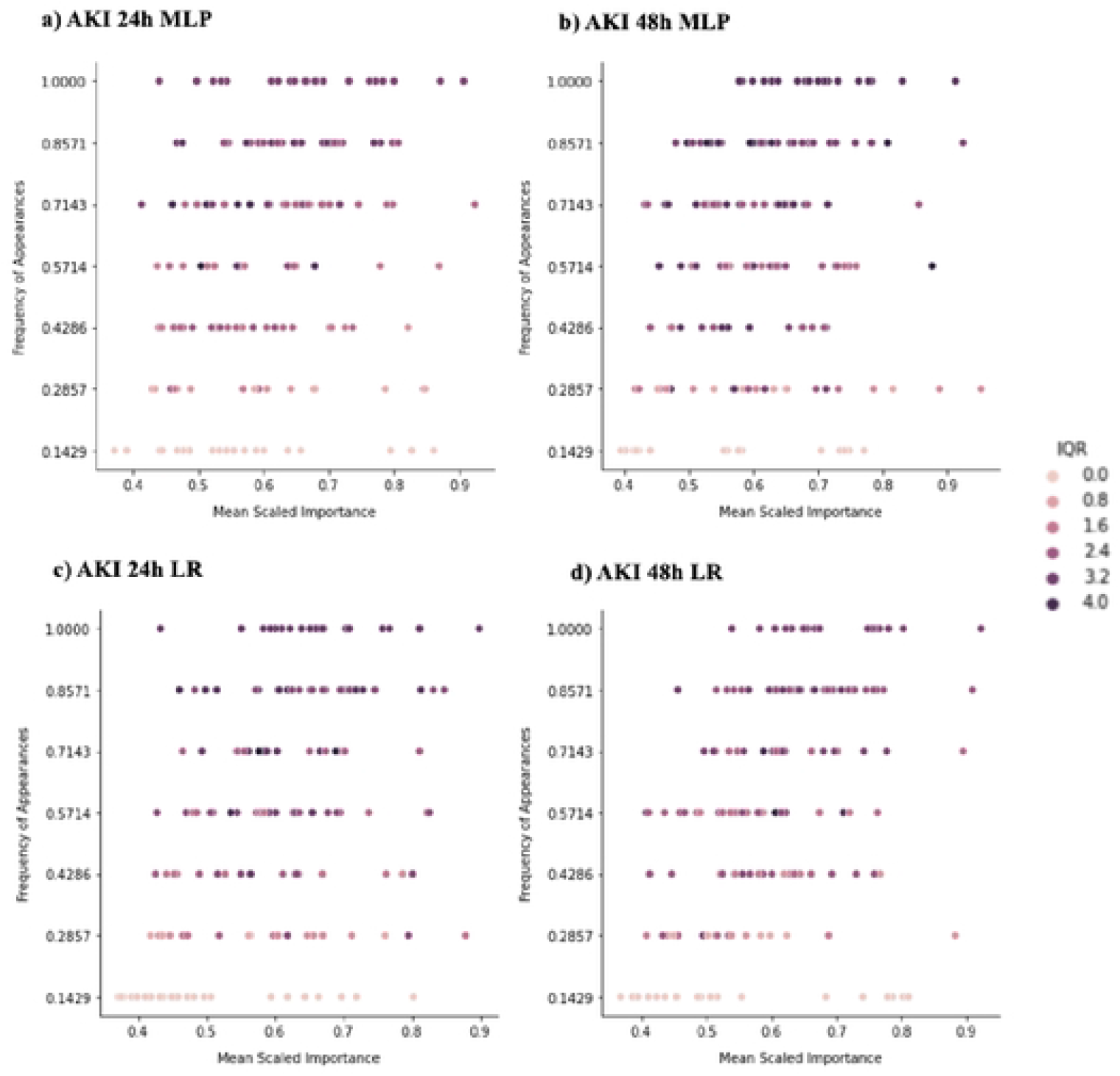
Distribution of important features at all local models across local sites. The figure demonstrates feature importance disparities for all AKI settings (24h. 48h) local models (MLP and LR). (a. b) show feature importance disparities for MLP models, (c. d) show feature importances for LR models. Each dot corresponds to one of the most important features ranked among the top-100 by at least one of the seven models: y-axis measures the proportions of sites that identified the feature as top-100, or “commonality across sites”; x-axis measures the mean of feature importance rankings measured as “soft ranking” (the closer it is to I. the higher the feature ranks). Top-100 is an arbitrary cutoff we used to analyze the most important features to illustrate heterogeneity. Each feature is also color coded by the interquartile range (IQR) of the ranks across sites (the higher the IQR is. the more disagreement across sites on the importance of that feature).

**Fig 6.**
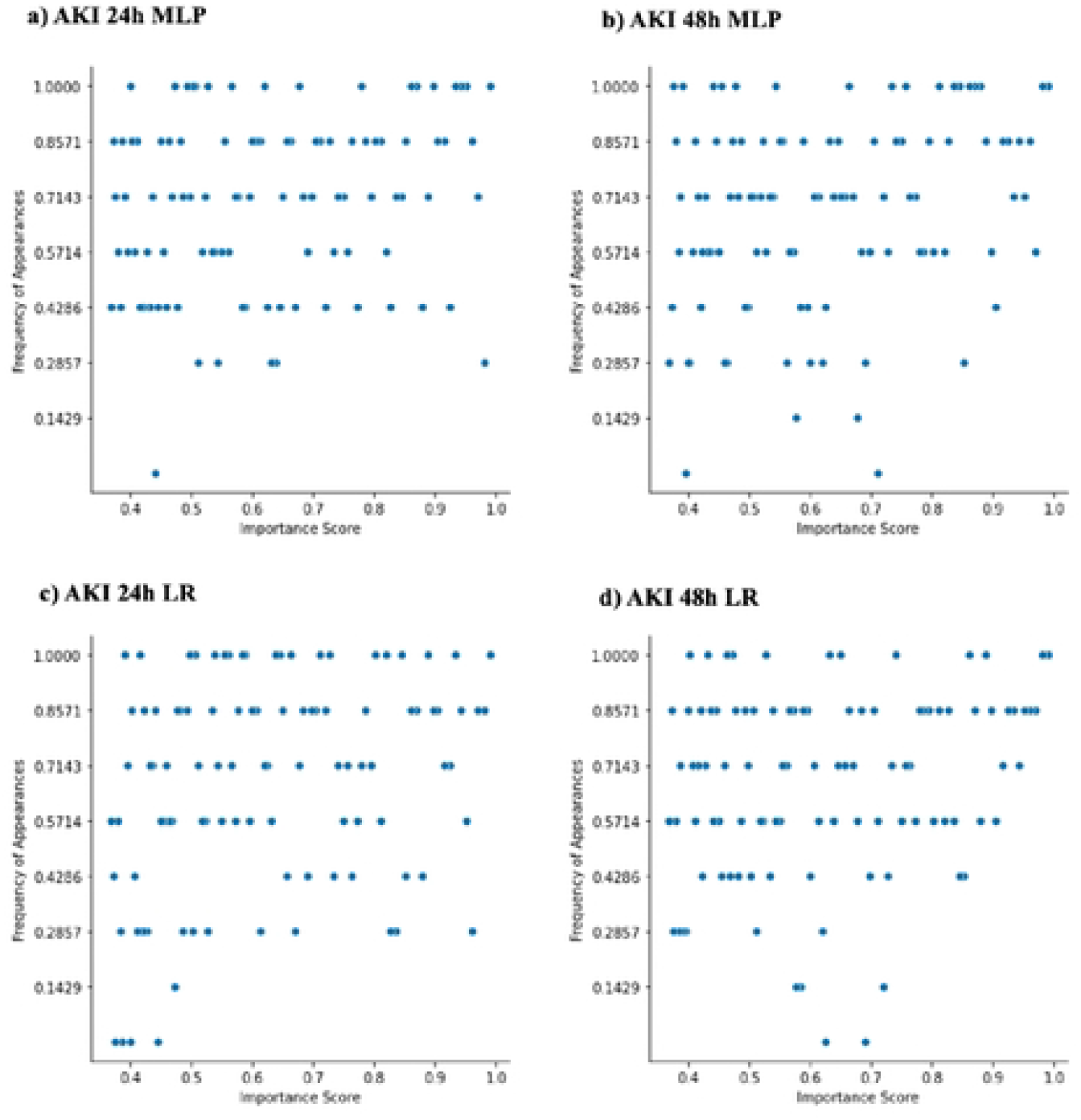
Distribution of important features at pooled models across local sites. The figure demonstrates feature importance disparities for all AKI settings (24h, 48h) pooled models (MLP and LR). Each dot corresponds to one of tire most important features ranked among the top-100 by the pooled model: y-axis measures the commonality across sites: x-axis measures the feature importance soft rankings.

**Fig 7.**
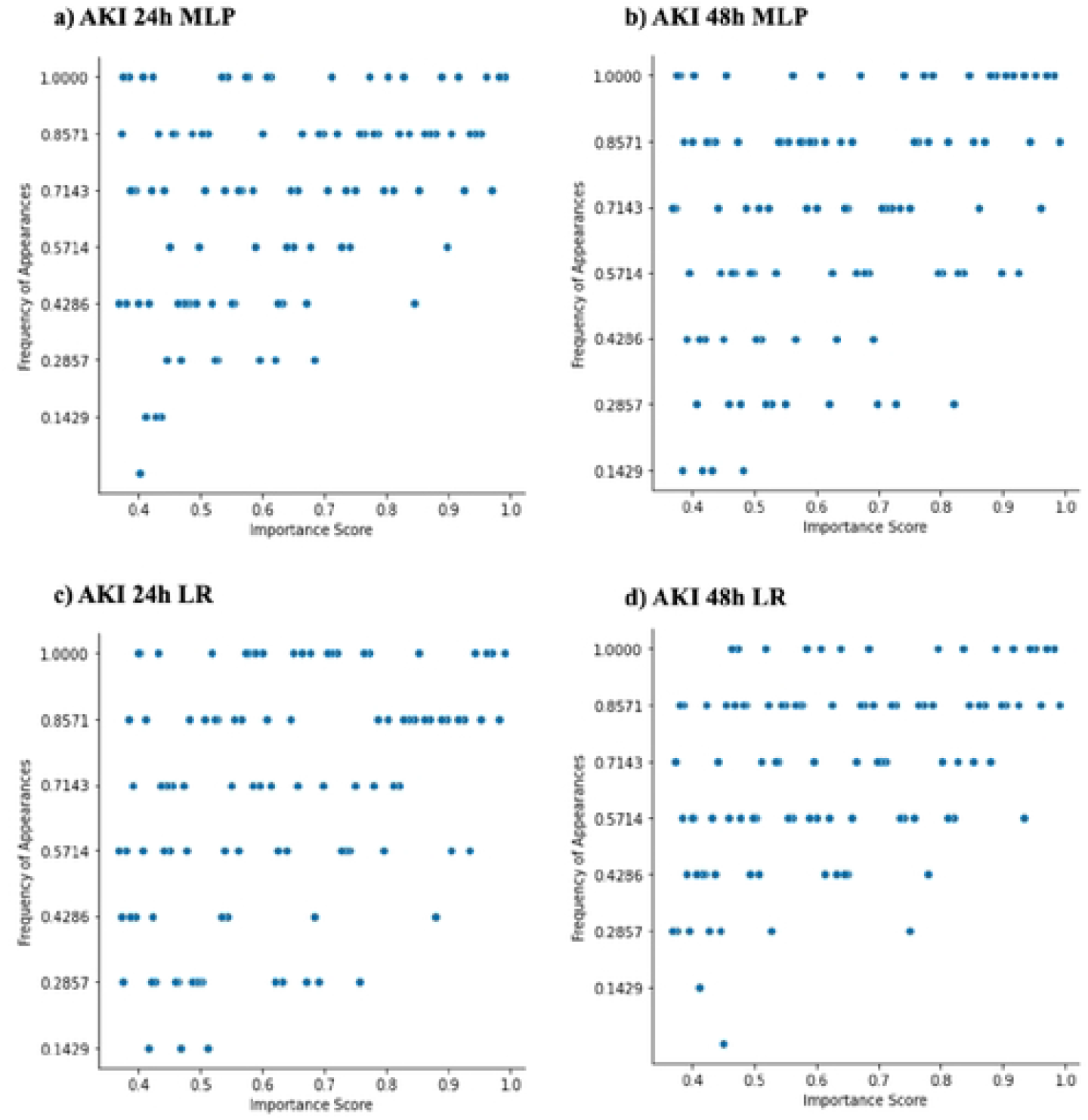
Distribution of important features at federated models across local sites. The figure demonstrates feature importance disparities for all AKI settings (2411. 48h) federated models (MLP and LR). Each dot corresponds to one of the most important features ranked among the top-100 by the pooled model: y-axis measures the commonality across sites: x-axis measures the feature importance soft rankings.

Fig 5 shows the distribution of important features across sites for local models. For all settings, there are features that are both ‘universally important’ and site-specific (i.e. important at only a subset of sites). The universally important features across most sites for AKI (both 24h and 48h) included *creatinine_last, creatinine_min, creatinine_max*, administration of Ondansteron, *glucose_max*, and *urineoutput_sum*. However, the relative importance of features at local sites were different. This disagreement was reflected in the dependence plots for the local models (S4 Fig). Universally important features like *creatinine_last*, administration of Sodium Chloride, among others, have different trends with the prediction diagnosis depending on the site.

Fig 6 shows the distribution of important features for the pooled models across sites. Both AKI 24h and 48h settings’ pooled models have relatively fewer features that are only important at a small number of sites compared to the local model framework. For the pooled MLP and LR AKI 24h or 48h model, the universally important features mainly included *creatinine_last, potassium_max*, and *creatinine_min*. All AKI pooled models have features that are uniquely important to the pooled models (i.e., these features were not considered as part of the top 100 features at any local site). The pooled MLP AKI 24h model uniquely considered administration of Nitroglycerin moderately important. The pooled LR AKI 24h model uniquely considered administration of Metoclopramide Mupirocin, Lidocaine, and *race_black* as slightly important. The pooled MLP AKI 48h model uniquely considered administration of Hydromorphone and *bilirubin_last* as slightly and moderately important respectively. The pooled LR AKI 48h model also uniquely considered administration of Hydromorphone and *bilirubin_last* as moderately important. Taken together, these differences suggested that there is slight variability in uniquely important features among models.

Fig 7 shows the distribution of important features for the federated models across sites. Similar to the pooled models, both MLP and LR federated models have relatively fewer features that are important at only a small number of sites compared to the local model analysis. The federated MLP AKI 24h model shares its universally important features with its pooled counterpart, namely attributing importance to *creatinine_last, potassium_max*, and *creatinine_min*, among others. These features are universally important in the federated LR architecture, as well as in the 48h setting. Some of the federated AKI models have uniquely important features as well. The federated MLP AKI 24h model considered administration of Dexmedetomidine as an important variable. The federated LR AKI 48h model considered administration of Phenylephrine slightly important. Similar to the pooled setting, we can see discrepancies between feature importances which are not considered universally important at all sites.

### Correlation of feature importances across model architectures

To investigate differences of feature importance between model architectures, we looked at the correlation between importance rankings of features shared by both the MLP and LR model for each setting and framework. Fig 8 shows these correlations, where the x and y axes are the importance of the feature in the MLP and LR model respectively. In the AKI 24h and AKI 48h (not shown) settings, local models have moderately strong positive correlations with PC ranging from 0.79 - 0.84. The pooled and federated AKI 24h model shows slightly weaker positive correlations as compared to the local models PC = 0.79, 0.77. These results suggest that, within the AKI setting, the pooled and federated models were not successful at decreasing feature discrepancies between the LR and MLP architectures that were present in local models. Sepsis prediction setting results are available in Supplemental Information and S9 Fig.

**Fig 8.**
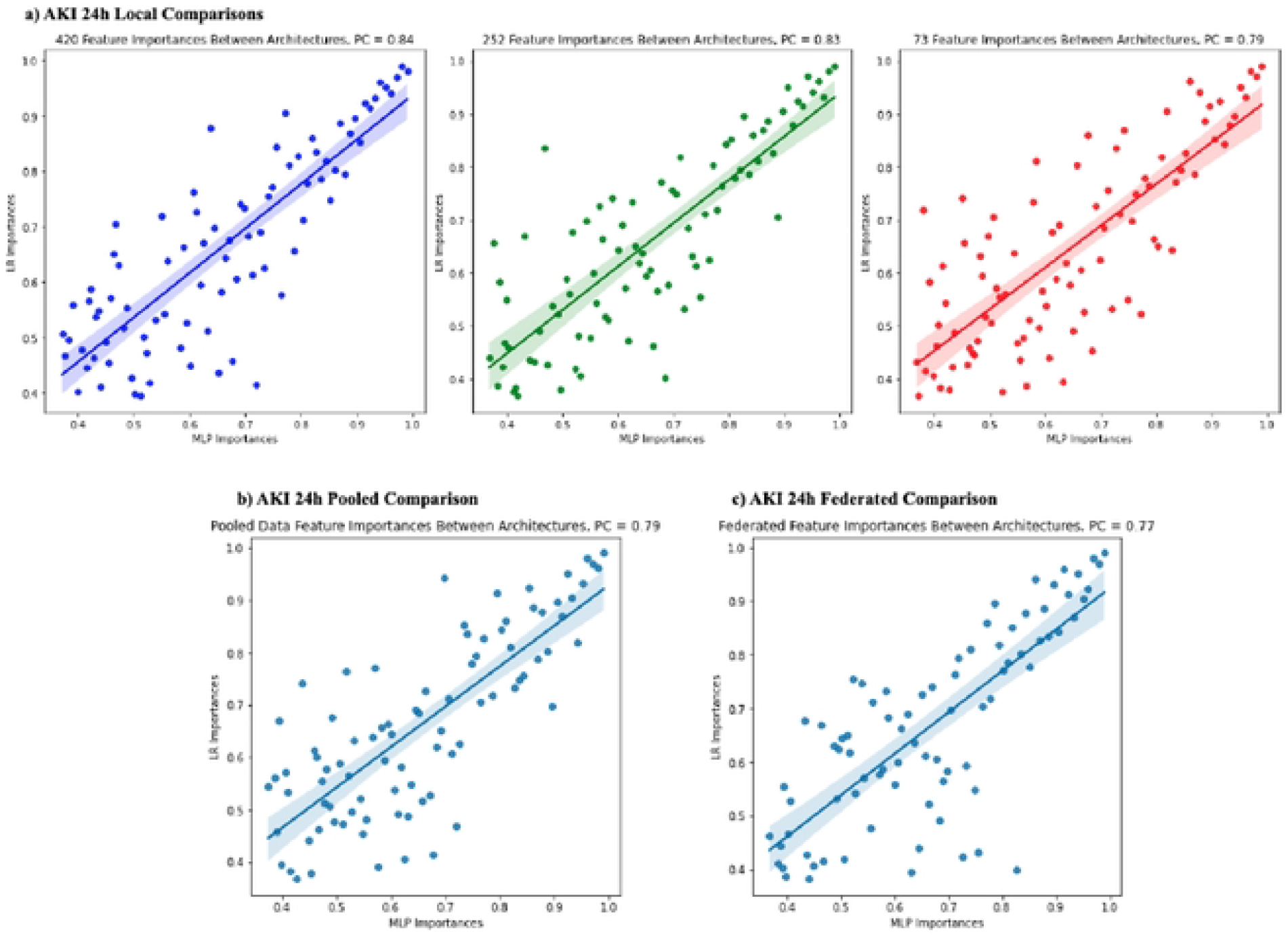
Important features comparison across model architectures for AKI 24h setting. The figure shows correlations between important features in the MLP and LR models. Each dot corresponds to one of the most important features ranked among the top-100 by both the MLP and LR model. The y-axis measures the importance of the feature in the LR model whereas the x-axis measures the importance in tire MLP model. The shaded portion represents a 95% confidence interval. PC (pearson correlation coefficient) for each comparison is denoted on the (op-left of each plot, (a) shows the comparisons for local sites, 420, 252, and 73. (b) shows comparisons for the pooled models, (c) show s comparisons for the federated models.

### Correlation between local feature importances and non-local framework feature importances

To investigate the correlation of heterogeneous features between local frameworks and both pooled and federated frameworks, we established the ‘unique importance score’ (UIS). The UIS score is large for features that are highly important at a small subset of sites, whereas it is small for features that are considered universally important (i.e. features that were important at a plurality of sites). In other words, the score is large for features that lie in the bottom right region of the plots in Fig 5, 6, and 7. Calculation of the UIS score can be found in the Methods section. Fig 9 shows the correlations of the UIS score across frameworks. Similar conclusions can be derived from both pooled and federated framework analyses, in both sepsis and AKI. There is a strong positive correlation (PC ranging from 0.84 - 0.93) between local UIS and pooled/federated. Interestingly, for all analyses, confidence on the line of best fit decreases at larger UIS scores. This suggests that features considered universally important in the local framework were important for the pooled/federated models whereas features only important at a small subset of hospitals were disregarded.

**Fig 9.**
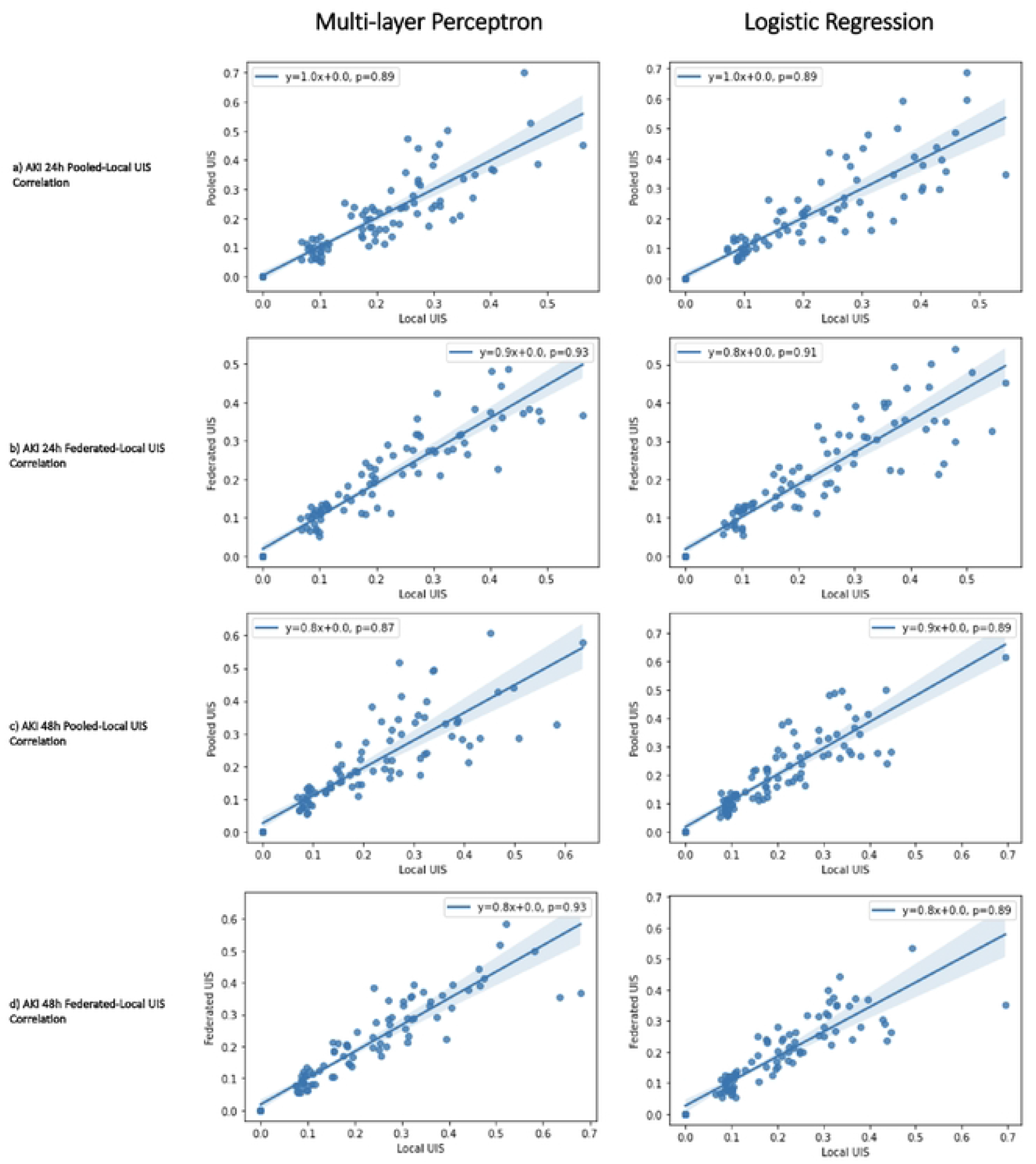
Correlation of unique importance .score (UIS) between local and pooled/federated frameworks. x-axis is the UIS for each feature in the local model framework, y-axis is the UIS for the pooled/federated model. Line of best fit is plotted and equation is shown on top comer, along with pearson correlation coefficient (p). Shaded area represents a 95% confidence interval. First column depicts plots for the MLP model whereas the second column depicts plots for the LR model, (a. b) show analyses for sepsis setting, (c. d) show analyses for AKI 24h setting, (a. c) show analyses for pooled framework, (b. d) show analyses for federated framework.

### Sources of data heterogeneity

Tables 1a and 1b show demographic profiles across each site for AKI and sepsis patients, respectively. For both AKI and sepsis settings, sites show similar gender distributions, with a slight majority of patients being male across all sites. Age distributions are also similar across all sites with a majority of patients being between 50 - 75 years old. Patient BMI is more or less similar across sites with most patients having a BMI between 23 - 34. Site 199 has slightly fewer patients with a BMI of less than 23, and more patients with a BMI of greater than 34 compared to other sites. In both settings, there was a disparity in the amount of patients that underwent elective surgery, with the proportions ranging from 0.12 - 0.28. Patients show differences in racial breakdowns across sites. The African American population varies across sites from 0.02/0.01 (AKI/sepsis) at Site 199 to 0.3/0.32 at Site 243. Site 73 has a relatively large population of Hispanic individuals compared to other sites, whereas Sites 122, 243, 252, and 458 have no Hispanic patients. The Asian population is similar across all sites. The ‘Other’ racial category has the largest proportion of individuals across all sites but this proportion varies largely depending on the site, ranging from 0.67 - 0.98. As previously mentioned, the majority of the patients in all settings (AKI 24h, 28h, and sepsis) were negative for the disease. For AKI 24h (not shown in Table 1a) and AKI 48h settings, the proportion of AKI positive patients ranges from 0.06/0.08 (24h/48h) to 0.1/0.13. For the sepsis setting, the proportion of positive patients ranges from 0.02 to 0.20.

**Table 1.**
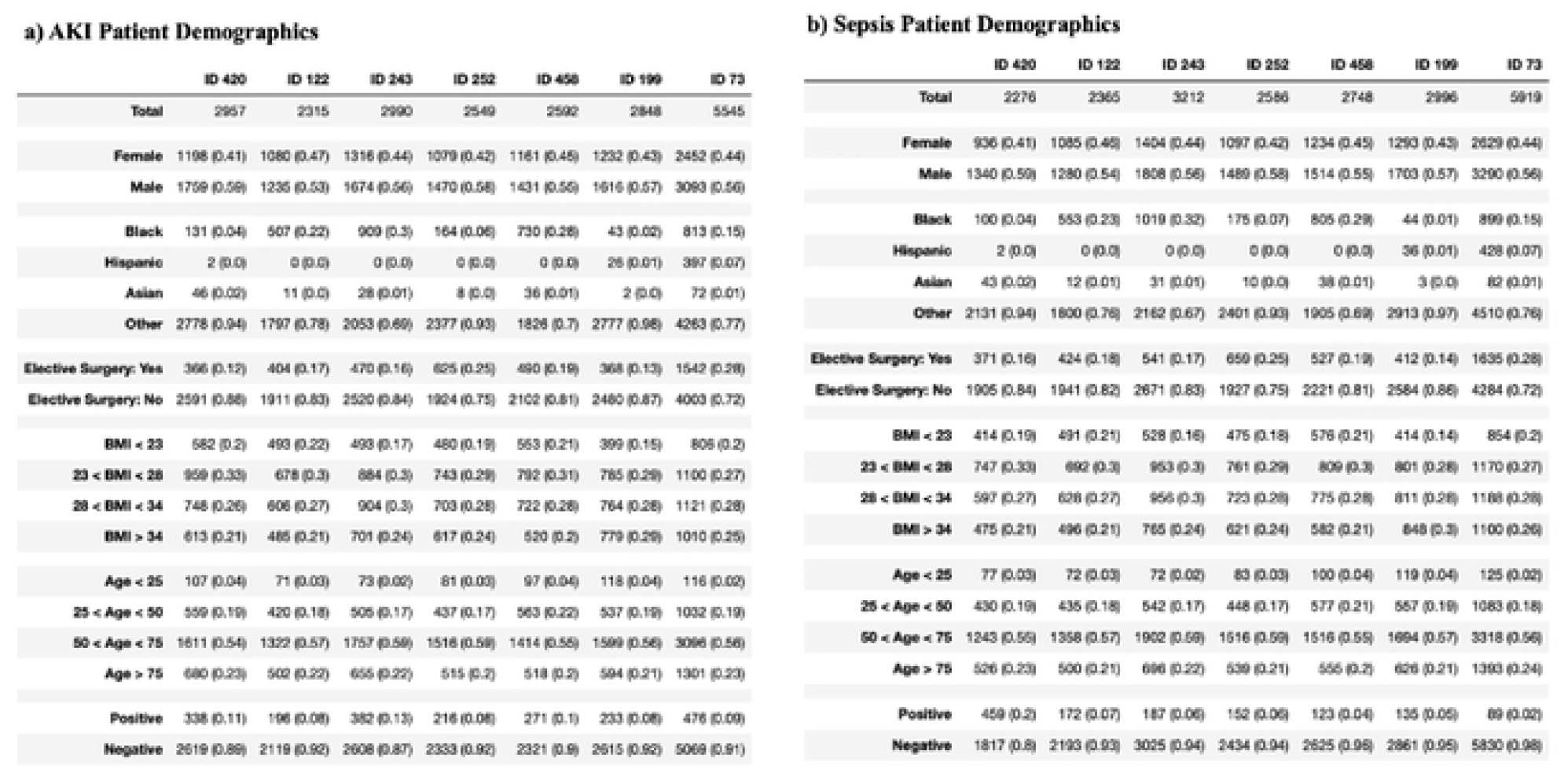
Demographic Characteristics of AKI (48h) (a) and Sepsis (h) patients at each site. Percentage of individuals with certain characteristics specified within parentheses.

Table 2 shows general site information for the 7 hospitals. The sites are located across the Northeast, Midwest, and South of the continental United States of America. All sites are large with greater than 500 beds. There are differences in patient unit types across all sites. Sites 420, 243, 252, and 199 have no patients in Cardiothoracic Intensive Care Units (CTICU). Sites 252 and 458 have no patients in Medical Surgery Units (Med-Surg ICUs). Sites 122 and 199 have no patients in Surgical Intensive Care Units (SICU). Sites 122, 243, 458, and 199 have no patients in Critical Care Cardiothoracic Intensive Units (CCU-CTICU). Sites 420 and 122 have no patients in Cardiothoracic Intensive Care Units (MICU). Sites 420, 122, and 199 have no patients in Neurological Care Units (Neuro ICU). Sites 252, 199, and 73 have no patients in Cardiac Intensive Care Units (Cardiac ICU). Among sites which do share patients in the same unit, proportions may be different. For example, while both Site 199 and 73 have patients in the Med-Surg ICUs, 88% of patients in Site 199 are admitted to Med-Surg ICUs whereas only 16% of patients in Site 73 are admitted to Med-Surg ICUs. Sites also had disparities in patient admission sources. Across hospitals, most patients were either admitted directly or admitted from the emergency department or operating Room. Of particular note, at Site 199, 22% of patients were admitted from the ICU to a special care unit (SCU). At Site 73, no patients were admitted from the recovery room. Taken together, despite being large, sites have disparities in unit types and sources of admission.

**Table 2.**
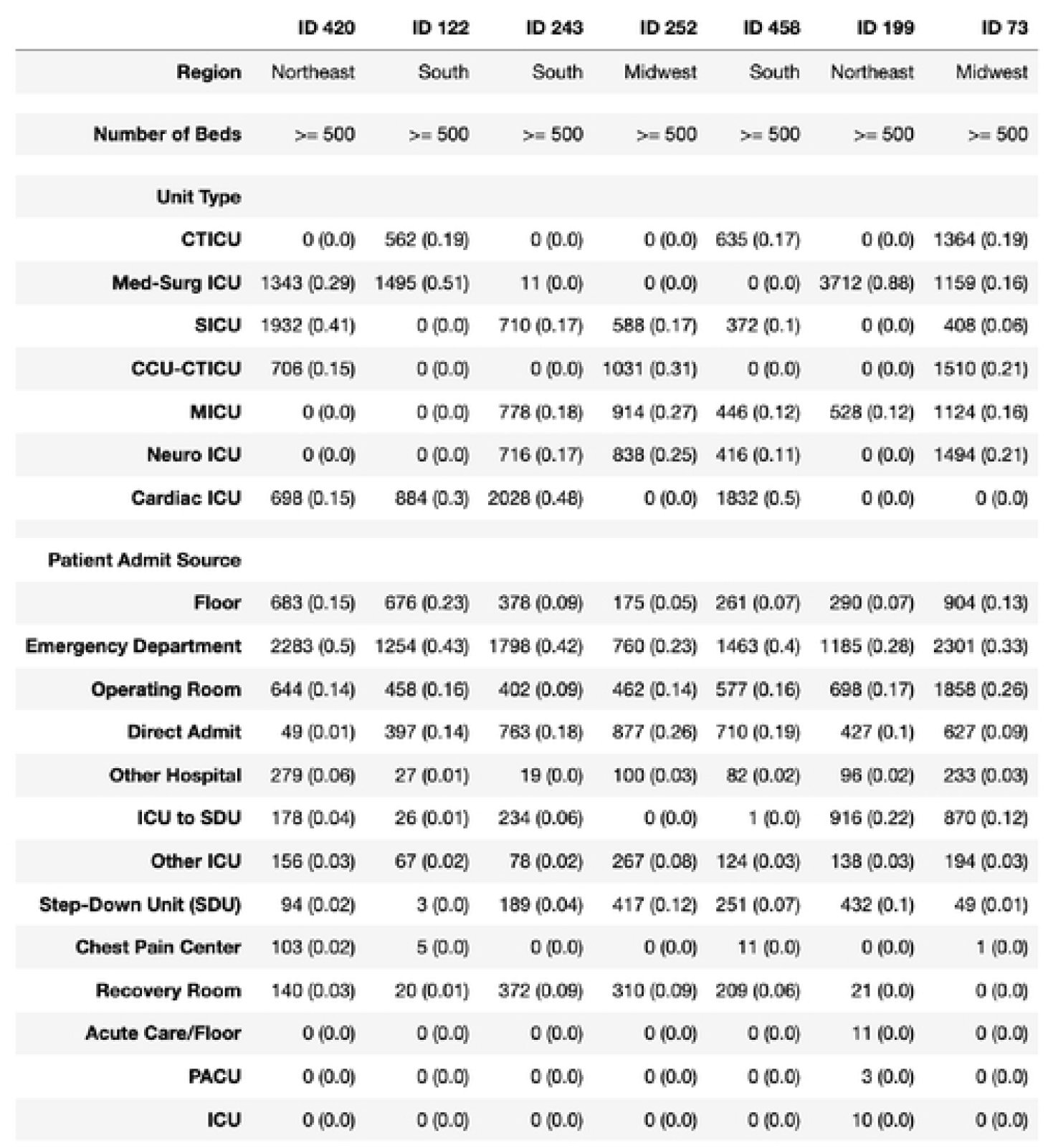
Hospital site information. Percentage of usage/individuals specified within parentheses.

S5 Fig illustrates the usage of medications across sites. Only 22 medications are used at all sites for both sepsis and AKI settings. Further analysis indicated that even for medications used at multiple hospitals, the proportion of patients that were on the medication at each hospital varied greatly. Coupled with the disparities in unit types, this suggests that each hospital site treats significantly different populations of individuals, despite all these hospitals having patients who suffer from AKI and sepsis.

## Discussion

In this study, to investigate the data heterogeneity in FL, multiple machine learning models were developed to predict the risk of both AKI and sepsis diseases in multiple ICU settings. Different types of EHRs including lab tests, vital signs, demographics, and medications were extracted from seven hospitals in the eICU collaborative research database. Three model frameworks including local, pooled, and federated were explored. Effects of data heterogeneity across hospital sites were evaluated through model performance comparison and feature importance analysis. The sources of data heterogeneity across hospitals were investigated based on patient demographics, medication usage, and general hospital attributes.

Our prediction models have shown comparable performance with state-of-the art AKI and sepsis prediction studies [7]. Federated model frameworks generally outperform their local counterparts, indicating that use of FL in creating AKI and sepsis prediction models trained on EHR data is warranted. The FL framework outperformed the local model framework, which was consistent with literature suggesting that FL can provide performance increases to sites where local model framework performance suffers [13]. Interestingly, the pooled model framework did not show much improvement compared to the local model framework. Based only on the performances of the different model frameworks, it seems that the presence of data heterogeneity may negatively affect pooled model framework performances whereas it may positively affect federated model framework performances.

The performance heterogeneity of predictive models across sites and frameworks was evaluated by comparing feature importance. For both AKI and sepsis prediction tasks, important variables identified by predictive models were consistent with prior studies [7]. For example, creatinine and furosemide exposure showed positive associations with AKI, which is unsurprising given their clinical association with AKI. For the same model architecture, importance of a feature varied depending on the sites, with variable-prediction relationships changing (see Fig 5, 6, 7). The presence of ‘universally important features’ (i.e., features that were considered highly important at a majority of sites) and ‘uniquely important features’ (i.e., features that were highly important at a small subset of sites) showed that there was disagreement on relative importance across sites. The feature heterogeneity plots for federated and pooled frameworks showed a decreased amount of uniquely important features. This was indicative of both these frameworks being able to attribute higher importance to features shared across multiple sites.

Our findings also demonstrated that federated and pooled models were not successful at decreasing feature importance discrepancies between LR and MLP architectures, and that both pooled and federated frameworks prioritized features that were considered important across a plurality of sites (i.e. low UIS) and attributed lower importance to features that were uniquely important at a small subset of sites (i.e. high UIS). These findings also suggest that the federated model may be better at discriminating the key features of patient-level clinical, lab, and demographic information that improves risk prediction. In the field of critical care medicine, the implication of this finding is that across heterogeneous sources of data, federated models are more likely to highlight the common elements that can better predict sepsis and AKI between hospital, patient, and practice-specific circumstances, thus highlighting the generalizability of the model’s value. However, as a consequence, it is possible that important local characteristics that may better predict AKI and sepsis within hospitals could be overlooked when compared to pooled or local models, which may in turn limit the clinical utility of these tools, a finding that is increasingly being acknowledged in the AI/ML literature.

Within our analysis, differences in features of the hospitals, and ICUs were notable. Many sites did not have any patients admitted to ICU types that other sites had a high proportion of patients within, for example the Medical Surgery ICU or SICU. At these hospitals, different sites treat different conditions. Thus, treatments may vary depending on the etiology and nature of the condition driving sepsis and AKI [14]. For example, a patient managed for decompensated heart failure in a cardiac ICU who subsequently develops AKI may be treated with inotropic support and furosemide, whereas a patient being managed for septic shock in a medical ICU with AKI may be aggressively repleted with intravenous fluids. As such different hospitals, which are specialized at treating different conditions, may have slightly differing medication regimens for treating patients when faced with the same disease, which in turn may be a function of the practicing physician and their choice of treatment options including medications, prespecified protocols, or even higher level decisions about cost within centralized hospital pharmacies. Our models highlighted this putative disagreement between medication usage at hospitals, creating another source of heterogeneity in model training. This heterogeneity in medications and demographic details was demonstrated in the feature heterogeneity plots since features with higher UIS scores tend to be medications and demographic information. Differently, lab tests and vital signs were generally universally important features across hospitals, likely because these are commonly standardized across hospitals. Taken together, local frameworks may heavily suffer in generalizability even when population demographics are similar across sites, due to disagreements in medication and treatment administration.

However, clinicians may find use in site-specific factors, which may not be evident in federated frameworks and only be ascertainable within a local framework. As such, while federated frameworks may provide performance increases, local frameworks can still provide clinical value in determining important site-specific factors for risk prediction.

## Limitations

There are several limitations to our study. First, we mainly considered structured clinical information to construct the features. Integrating unstructured free text to build predictive models may obtain better predictive performance and allow a new level of explainability of prediction. Second, we only considered LR and MLP to build predictive models based on local, pooled, and federated frameworks. Other algorithmic solutions such as support vector machines may have a potential to improve model performance. Third, we mainly focus on describing the effects of data heterogeneity in FL in terms of disease risk prediction, considering data harmonization techniques to mitigate the problem and improve the performance is one of future research topics.

## Data Availability

All data are in the manuscript and/or supporting information files. Scripts for processing and analyzing data are available at https://github.com/surajraj99/Data-Heterogeneity-in-Federated-Learning.

https://eicu-crd.mit.edu/

## Acknowledgements

SR would like to acknowledge the support from Tri-Institutional Training Program in Computational Biology and Medicine (CBM) funded by the NIH grant 1T32GM083937. ZX, WP, FW would like to acknowledge the support from NSF 1750326, NSF 2212175, NIH R01AG076234, NIH RF1AG072449, Google Faculty Research Award and Amazon Machine Learning Research Award.

## Methods

### Data Aggregation

Patient data was extracted from the eICU Collaborative Research Database, a multi-center critical care database made publically available through Philips Healthcare and the MIT Laboratory for Computational Physiology (https://eicu-crd.mit.edu/). The database contains detailed information regarding the clinical care of ICU patients. We investigated three disease settings (24h or 48h observation window (OW) AKI, and sepsis). An AKI (non-graded) is defined as any of the following: Increase in serum creatinine (SCr) by >=0.3 mg/dl (>=26.5 μmol/l) within 48 hours, increase in SCr to >=1.5 times baseline which is known or presumed to have occurred within the prior 7 days, or urine volume < 0.5 ml/kg/h for 6 hours. We predict AKI risk using an accumulating OW (S1 Fig). We predicted AKI within the next 24 hours (prediction window, PW) following the end of the OW, focusing only on the first 3 days (72 hours) of a patients’ inpatient hospital stay (max OW = 48 hours). For each patient, we created 2 pairs of OW/PWs, specifically using from OW=1-24 hours (1-day) after admission, 1-48 hours (2-days). We do not consider the onset point. For the AKI prediction experimental setting, positive cases are samples that are diagnosed as AKI in the prediction window whereas controls are samples that are not diagnosed as AKI in the prediction window. For sepsis prediction, we labeled patient data in accordance with Sepsis-3 clinical criteria. We predicted whether patients will suffer from sepsis 6 hours prior to onset, onset point included. For sepsis prediction experimental setting, positive cases are samples that are diagnosed as sepsis. Controls are samples that are not diagnosed as sepsis. For patients who did not develop sepsis, predictor values were selected from a random T-hour time window (T is usually set as 48 or 24 hours) during the patient’s ICU stay. For those who developed sepsis, a time was selected for the patient within admission to 6 hours prior to the onset of sepsis, and the predictor values were extracted. Data was collected from seven hospitals with the following IDs: 420, 122, 243, 252, 458, 199, and 73. For all three disease predictions (24h or 48h AKI, and sepsis), all hospital sites shared all features including: general demographic information (8 variables), vital signs/lab tests (29 variables), and medications (254 medications). For 28 vital signs and lab tests, the max, min, first, and last values are calculated. For urine, only the summation is calculated. Taken together, a total of 354 features were available at every hospital site for each patient.

### Data Processing

For all datasets, we performed an automated curation process outlined as follows: (1) systematically identified extreme values of numerical features (e.g. vital signs/lab tests and some demographic information) that were beyond the 1st and 99th percentile as outliers. We marked these values as missing. (2) We standardized all our variables appropriately by normalizing all our numerical features and converting binary features to either 1 or -1. (3) For all missing measurements, the Multiple Imputation by Chained Equations algorithm (MICE) was used. MICE imputation is able to calculate missing information by taking advantage of the relationships between non-missing measurements within the dataset.

### Experimental Design

There were three prediction tasks including 24-hour and 48-hour prediction of AKI, and sepsis prediction. Three model frameworks were designed including local, pooled, and federated model frameworks. The local model framework only used data from each site itself. The pooled model framework combined data from all sites. In the federated model framework, each local site does not have access to other sites’ data. A model was trained locally and its parameters were shared to a central aggregator, which was used to update global model parameters which were subsequently sent back to each site. For each framework, LR and MLP were used as model architectures, so there are 54 tasks in total were performed (7 site-specific (local) x 3 prediction tasks x 2 architectures + 1 pooled model x 3 prediction tasks x 2 architectures + 1 federated model x 3 prediction tasks x 2 architectures). For all settings, five fold cross-validation was used during training models. The Shapely Additive exPlanations (SHAP) tool was used to calculate feature importance rankings for each task. The Markov Chain Type 4 rank aggregation was used to combine the feature importance rankings for all five folds.

### Learning Algorithm

We chose Multilayer Perceptron as the learning model. A multilayer perceptron (MLP) is a class of feedforward artificial neural network (ANN). An MLP consists of at least three layers of nodes: an input layer, a hidden layer and an output layer. Except for the input nodes, each node is a neuron that uses a nonlinear activation function. MLP utilizes a supervised learning technique called backpropagation for training. Its multiple layers and non-linear activation distinguish MLP from a linear perceptron. It can distinguish data that is not linearly separable. The MLP consists of three or more layers (an input and an output layer with one or more hidden layers) of nonlinearly-activating nodes. Since MLPs are fully connected, each node in one layer connects with a certain weight to every node in the following layer. To implement the MLP model, Python’s PyTorch library was used. PyTorch is an open source machine learning framework based on the Torch library, used for applications such as computer vision and natural language processing, primarily developed by Facebook’s AI Research lab. All MLP models had one hidden dense layer of 10 units, learning rate = 0.001, used binary cross-entropy loss, and stochastic gradient descent optimization. To mitigate class imbalance, class weights were used to penalize the loss for positive class inaccuracies. Each model was trained for 200 epochs and the batch size was 64.

In addition to the Multilayer Perceptron model, we implemented a Logistic Regression (LR) model. The LR model consisted of one linear layer followed by sigmoid activation. Similar to the MLP, a learning rate = 0.001, binary cross-entropy loss, and stochastic gradient descent optimization was used. Class weights were applied in a similar fashion to the MLP model. For consistency and to enable direct comparisons, all models of each framework for all tasks were built with the same architecture.

Our primary model framework of interest was a federated learning model. In this model, training was performed in different sites, and parameters were shared to a central location. To create a federated model using both the MLP and LR architecture, the federated averaging technique was used. The process was as follows: a central aggregator initialized the federated model with random parameters. This model was sent to each site, then trained for one epoch. Next, model parameters were sent back to the central aggregator where federated averaging was performed. Updated parameters from the central aggregator were then sent back to each site, and this cycle was repeated for multiple epochs. Federated averaging scales the parameters of each site according to the number of available data points and sums all parameters by layer. Through this technique, federated models did not receive any raw data. Class weighting was performed at each site on every cycle, which ensured local data distribution information was not sent to the global server. All parameters for local server models were kept the same to enable comparison. We were able to perform federated class weighting through this mechanism because local data distributions were similar across hospitals.

### Evaluations

We used the area under the receiver operator curve (AUROC) to compare the overall prediction performance, which is known to be more robust to imbalanced datasets. In addition to AUROC, accuracy, precision, and recall were calculated. In addition to aggregate performance metrics for each model, training loss and training/testing AUROC histories were measured. Tests of significance were performed using the Student’s t-test. Feature importance rankings for each task were computed using SHAP. To focus more on the most impactful features (i.e., variables ranked among top 100) without losing information on the weaker features, we assigned a”soft”membership of a feature as how high up the rank is relative to tops (s=100) by applying an exponentially decreasing function to the original ranks (r), i.e., f(r)=exp{−r/s}. For some top features, SHAP dependence plots were generated to illustrate the effect that each feature has on the predictions made by the model. Locally Weighted Scatterplot Smoothing (LOWESS) was used to fit a smooth trend line to the dependence plots.

The unique importance score (UIS) was calculated for each model architecture-setting-framework combination. For local model analysis, the mean importance *i*_*lj*_ for each feature *j* was calculated by averaging all soft rankings for said feature across all sites. This was done for all top 100 features at each local site. For both pooled and federated analysis, importance (*i*_*pj*_ or *i*_*fj*_) of each feature *j* was set to the soft ranking of said feature within the pooled or federated model. In all model frameworks, the frequency *f* of each feature was calculated by determining the proportion of local sites the feature was a top 100 feature. Given *i*_*lj*_, *i*_*pj*_, *i*_*fj*_, and *f*:

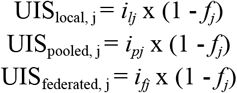

